# HIV profile of Neonatal Centre of Excellence Unit Admissions at the University Teaching Hospital-Children Division, Lusaka, Zambia after 2 decades of PMTCT: Retrospective pilot study

**DOI:** 10.64898/2026.01.24.26344762

**Authors:** Adenike Oluwakemi Ogah, James-Aaron Ogbole Ogah, Edwin Kanombola Chembo

## Abstract

**Objectives:** The aim of this study was to examine the prevalence of congenital HIV infection at the Neonatal Centre of Excellence Unit (NCOE), Children Division of the University Teaching Hospitals (UTH), and to analyze the characteristics of neonates who tested positive and negative for HIV PCR.

**Subject and methods:** This study is a pilot investigation that analyzed retrospective cross-sectional data from 757 mother-neonate pairs. The data was collected over a 12-month period from the ward register and file records at the NCOE Unit, UTH - Children’s Division in Lusaka, Zambia. The prevalence and characteristics of HIV among hospitalized neonates were assessed using percentages, Chi-square tests, ANOVA, and binary logistic regression model. The results were reported in terms of p-values, odds ratios, and 95% confidence intervals.

**Results:** In 2024, the annual rates of HIV and Syphilis among all neonatal admissions at NCOE were recorded at 4.1% and 5.9%, respectively. The HIV status for 52 neonates (6.9%) was not available. The median age of the neonates at the time of admission was 14 days, with an interquartile range (IQR) of 9 to 21 days. The maternal HIV positivity rate was 11.9%, while the paternal rate was 7.3%. Notably, a greater number and percentage of fathers had unknown HIV status (103, 13.6%) compared to mothers (20, 2.6%). The rate of mother-to-child transmission of HIV was observed to be 51.7%, while the rate of HIV exposure was 48.3%. A total of twenty-nine mothers, accounting for 3.8%, did not attend the antenatal clinic during their pregnancy.

Overall, the incidence of teenage pregnancies was 4.9%, and 47 mothers, or 6.2%, delivered outside of healthcare facilities. The rate of cesarean sections was 20.6%, and 57.3% mothers experienced delays in starting breastfeeding. Furthermore, 82.2% of neonates were referred from other healthcare facilities, and 73.6% showed indicators of growth faltering. A significant number of neonates presented at admission with abnormal body temperatures (60.1%), heart rates (66.8%), and respiratory rates (83%).

The characteristics of neonates diagnosed with HIV were comparable in all examined aspects to those without HIV. The sole distinctions observed were that mothers of HIV-positive neonates were, on average, significantly older (31.1 years, with a standard deviation of 5.40 years, representing a 3.37-year increase, and a 95% CI of 1.19 to 5.55, with a p-value of 0.003). Furthermore, these HIV-positive neonates had a greater propensity to be born to discordant couples (15.4% vs 1.5%; OR=11.72. 95% CI 3.35, 40.99; p=0.001); mothers with moderate parity (OR = 2.51; 95% CI: 1.05 to 5.88; p=0.032), to be born prematurely (OR=5.83; 95% CI: 1.15 to 29.58; p=0.047), and exhibited significantly impaired postnatal growth (OR= 3.02; 95% CI: 1.26 to 7.24; p=0.010) when contrasted with HIV-negative neonates. Notably, the impairment of postnatal growth manifested earlier than anticipated, with a substantial rate of 74.1% observed among the HIV-positive infants.

Neonates whose record of HIV status was missing presented with distinct characteristics compared to those with a known HIV status. Specifically, neonates with unknown HIV status were more frequently born to parents who were either HIV positive or whose HIV status was also unknown, as opposed to parents who were HIV negative (p<0.001). Furthermore, these neonates were less likely to have received antenatal care (p<0.001; OR=4.83, 95% CI 1.96-11.91). They also exhibited delayed initiation of breastfeeding, demonstrated impaired growth, and presented with relatively elevated random blood glucose levels and irregular body temperature at the time of admission.

The prevalence of discordant couples was observed to be 20.9% (19 out of 91couples). This rate was notably higher among infants whose HIV status was unknown, at 27.3% (6 out of 22), compared to 20% (10 out of 50) for infants with known HIV status, a statistically significant difference (p<0.001) with an odds ratio of 9.07 (95% confidence interval: 3.16 to 26.00). Among all the 19 identified discordant couples, 18 mothers tested positive for HIV, while only 1 father tested positive.

**Conclusion:** The significant number of mothers lacking antenatal care in this study is a cause for concern and poses a risk to the advancement of the Prevention of Mother-to-Child Transmission (PMTCT) program within the nation. Intensified antenatal care initiatives, encompassing early HIV screening for expectant mothers and their partners, are imperative to enable the timely initiation of Antiretroviral Therapy (ART) and PMTCT services. Particular vigilance is warranted for neonates presenting with an unknown HIV status in healthcare facilities within resource-limited environments. Such infants, especially those born to mothers aged >25years, with inadequate or absent antenatal care, those with moderate parity, premature births, delayed breastfeeding initiation, faltering growth, and abnormal vital signs, should be suspected of HIV positivity and undergo early infant testing to further mitigate infant morbidity and mortality. Regular assessment of these infants’ feeding, health, and growth shortly after birth is crucial, either through home visits or postnatal clinic appointments. Targeted counseling for mothers (and partners) with unknown HIV status, HIV positive infants, infants with unknown HIV status, women aged >25 years, belonging to discordant couples; and public education are essential for reducing HIV incidence and improving infant health outcomes within the community.

## Background

The HIV program’s prevention of mother-to-child transmission (PMTCT) is a public health intervention aimed primarily at reducing HIV transmission from mother to child by providing antiretroviral treatment (ART) to HIV-infected pregnant/postpartum women and prophylaxis treatment to exposed babies (Chiya et al., 2018)^1^. Although some countries in sub-Saharan Africa have achieved low rates of HIV mother-to-child transmission (MTCT), others, notably Zambia, continue to lag behind (Gumede-Moyo et al., 2019)^2^.

By the end of 2001, approximately 150,000 children in Zambia were living with HIV. About 90% of these infections occurred due to mother-to-child transmission. In the absence of intervention, more than 30% of these mothers would transmit the virus to their newborns each year. To address this problem, Zambia’s Ministry of Health established the PMTCT program in January 1999, Kankasa et al. (2002)^3^. Between 2010 and 2016, PMTCT programs have led to a 47% decline in infant HIV infections globally.

The components of Zambia’s PMTCT program are structured around four principal pillars or “prongs”: targeted interventions, including information, education, and counseling for young women and expectant mothers to prevent new HIV infections; community involvement and follow-up by employing community-based behavior change communication to promote safer sexual practices, reduce loss to follow-up, and enhance adherence to antiretroviral therapy; prevention of unintended pregnancies through family planning services; integration and linkage of reproductive health services with Child Health, tuberculosis services, and HIV testing and care within PMTCT; provider-initiated testing and counseling (PICT), implementing opt-out or routine HIV counseling and testing for all pregnant women during antenatal care, Mwanza et al.2022^4^.

In accordance with WHO guidelines, Zambia adopted Option B+, initiating lifelong triple-drug ART for all pregnant and breastfeeding women living with HIV regardless of CD4 count or clinical status. HIV-exposed infants receive ARV prophylaxis from birth for six weeks or throughout the breastfeeding period. Facility-based deliveries are promoted with skilled birth attendants, while procedures that elevate transmission risk are avoided. Counseling and support are provided on safe infant feeding practices, with exclusive breastfeeding recommended for the first six months, followed by continued breastfeeding alongside appropriate complementary foods. Early Infant Diagnosis (EID) involves HIV DNA-PCR testing of infants, typically at six weeks of age. HIV-positive mothers and their children are enrolled in long-term HIV care and treatment services. Psychosocial and nutritional support groups for mothers are offered to enhance retention in care, Hairston et al.2012^5^.

In 2015, Cuba achieved the elimination of perinatal HIV and syphilis (WHO 2015)^6^, whereas Zambia had set a target to attain HIV epidemic control by 2021. However, this objective remains unmet owing to challenges at multiple levels. The present study seeks to offer additional reflection on the performance of PMTCT as observed at one of Zambia’s eight Neonatal Centres of Excellence.

## Materials and methods

### Study Design

This is a pilot study, involving retrospective cross-sectional data at the NCOE unit of the University Teaching Hospitals-Children Division, Lusaka Zambia. Data on 758 neonates over a period of 12months were compiled during a ward audit in preparation for the main study investigating neonatal admissions for sepsis-Meningitis.

### Study setting

This pilot study was conducted at the NCOE unit located at the University Teaching Hospital-Children Division (a National Referral centre). NCOE is a 23 cot, level 2 Nursery catering for neonates admitted through the Emergency unit and general outpatient clinic.

### Data source and sampling method

Information on all neonates admitted consecutively between 27^th^ February 2024 and 5^th^ March 2025 was obtained from the NCOE and UTH-Child registry and file records. The results of the newborn blood smear cards for HIV DNA-PCR, maternal and partner HIV status, syphilis RPR status and socio-demographic data were obtained from the records in the file. The dependent variable was the HIV status of the newborn, which was recorded as either positive or negative. Independent variables were neonates’ syphilis and parents’ HIV status, birth weight, current weight, chronological age, gestational age, vitals (heart rate, respiratory rate, body temperature, oxygen saturation), number of weeks of gestation, place and mode of delivery, parental age and parity and ARV therapy.

### Statistical analysis

Data was manually cleansed, processed, verified for completeness, and entered into Microsoft Excel. The data were then exported to version 26 of the SPSS for analysis. After the categorization and definition of variables, descriptive analyses of each variable were performed, presented in numbers, frequencies and percentages. The chi-t test (Fisher’s exact test for small cells), One way ANOVA (Kruskal Wallis test for non-parametric data), Independent sample T-test (Mann-Whitney U test for non-parametric data) and binary logistic regression analysis were used to evaluate the characteristics of HIV-positive neonates. The multicollinearity and robustness of the regression model was verified. The regression model included factors with p values <0.1. The odds ratio (OR) was calculated with a 95-percent confidence interval (CI). Statistical significance was declared for all with a p value <0.05. The reporting in this study followed the STROBE Observational Guidance.^7^

### Ethics

The University of Zambia’s Institutional Review Board (UNZABREC) granted approval for the principal investigation into Neonatal Sepsis Meningitis, under reference number 5664-2024. The objective of this present study was to conduct an audit of ward admissions in anticipation of the aforementioned primary research. All documentation pertinent to the study was maintained securely within a locked cabinet. To safeguard the privacy and confidentiality of all participants, no personal identifiers, such as names, were collected. The findings derived from this study will be disseminated to all relevant stakeholders, serving as valuable information to inform strategies aimed at enhancing perinatal care.

## Results

### Participants

Over the course of 12 months, 758 mother-baby pairs were admitted to UTH-Children Hospital’s Neonatal Centre of Excellence. One baby’s record was excessively incomplete and was therefore excluded from the data. Data from 757 neonates were consequently analyzed.

### Characteristics of participants in the study

Univariate analysis revealed that 31 (4.1%) and 45 (5.9%) of the 757 neonates recruited tested positive for HIV and Syphilis, respectively, as shown in Table 1. The HIV status of 52 (6.9%) infants remained unclear. Two of the 47 (6.2%) neonates born outside of a health facility were noted: one was born on the way to the hospital and the other on a farm.

**Table 1:**
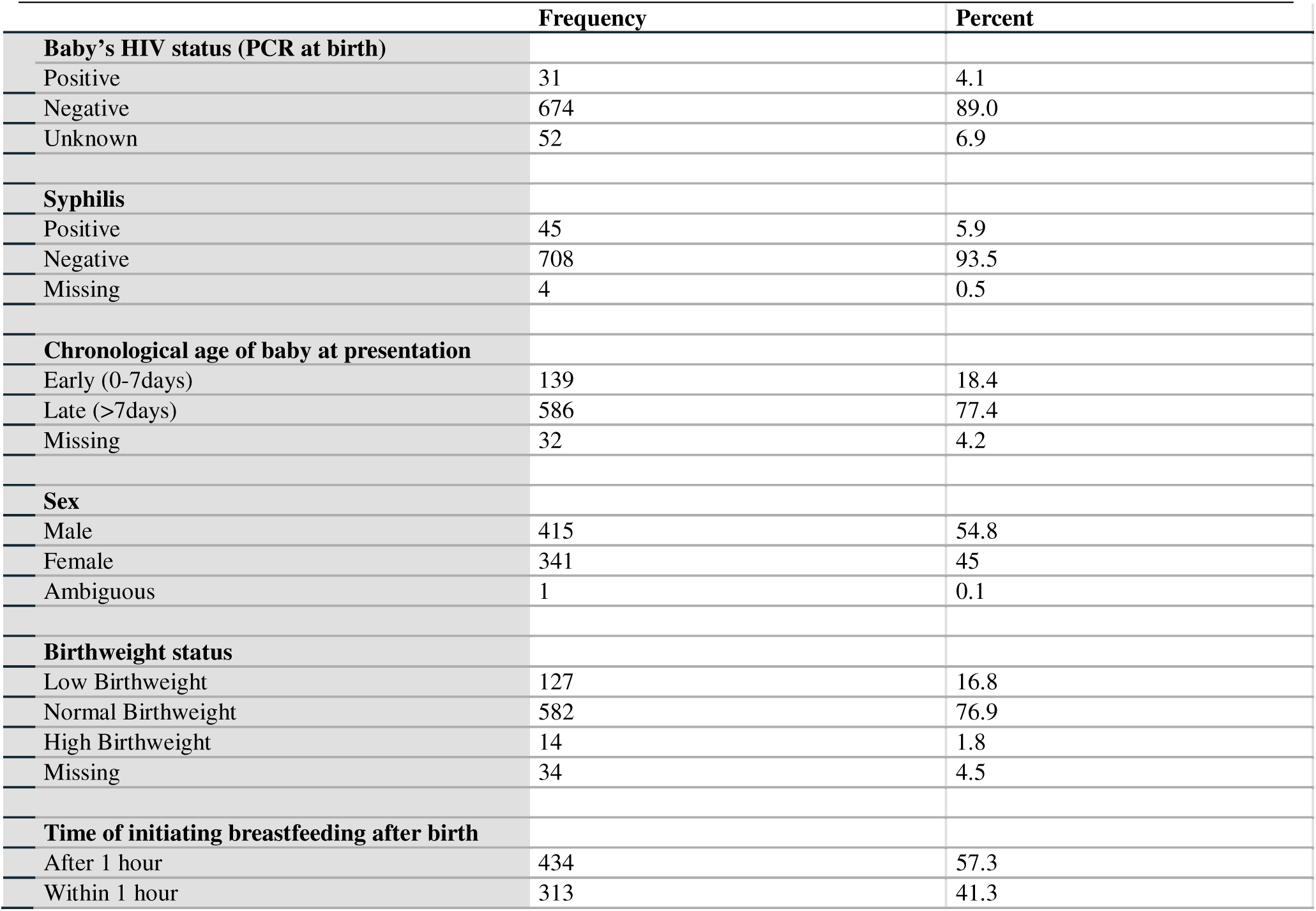
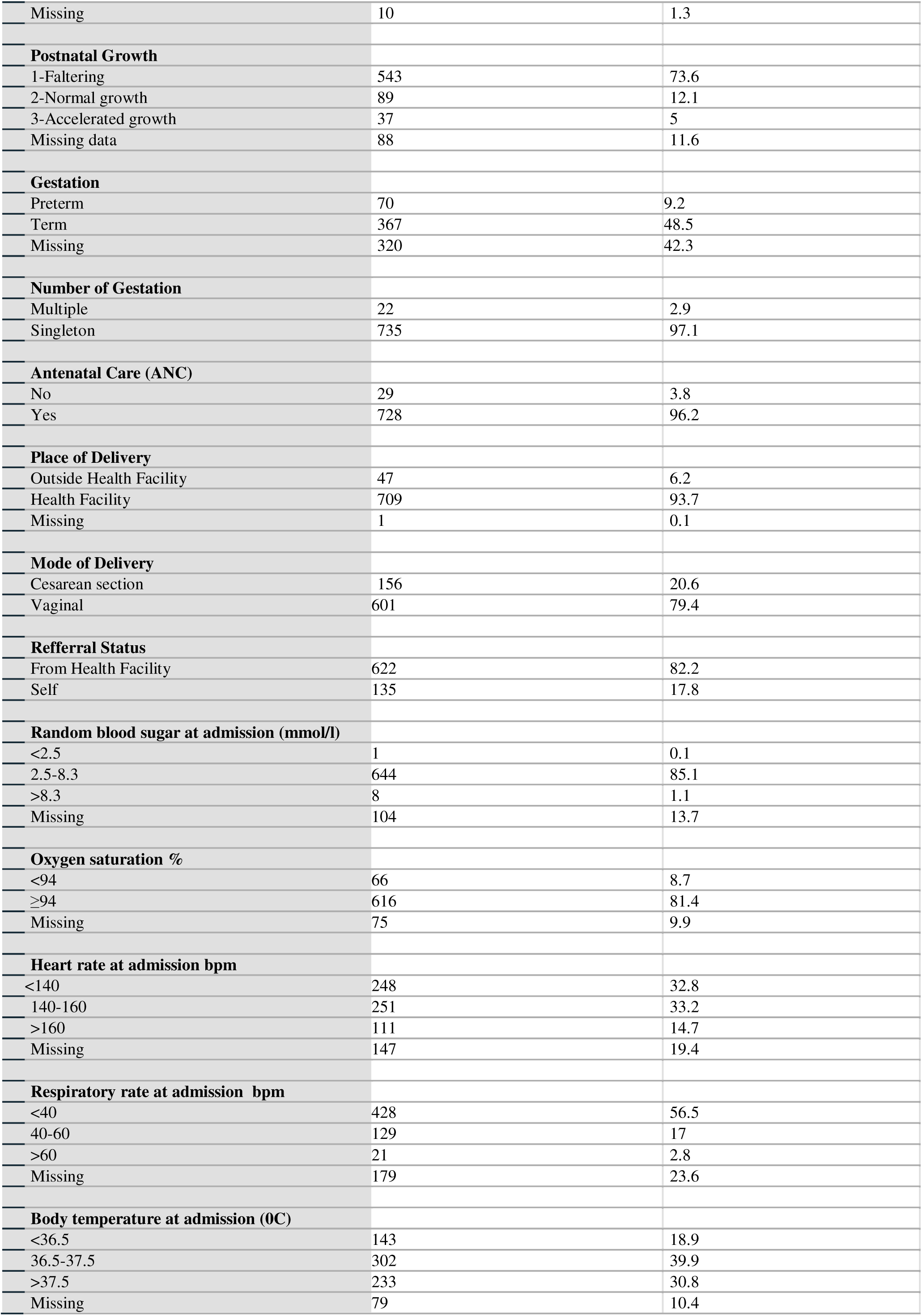
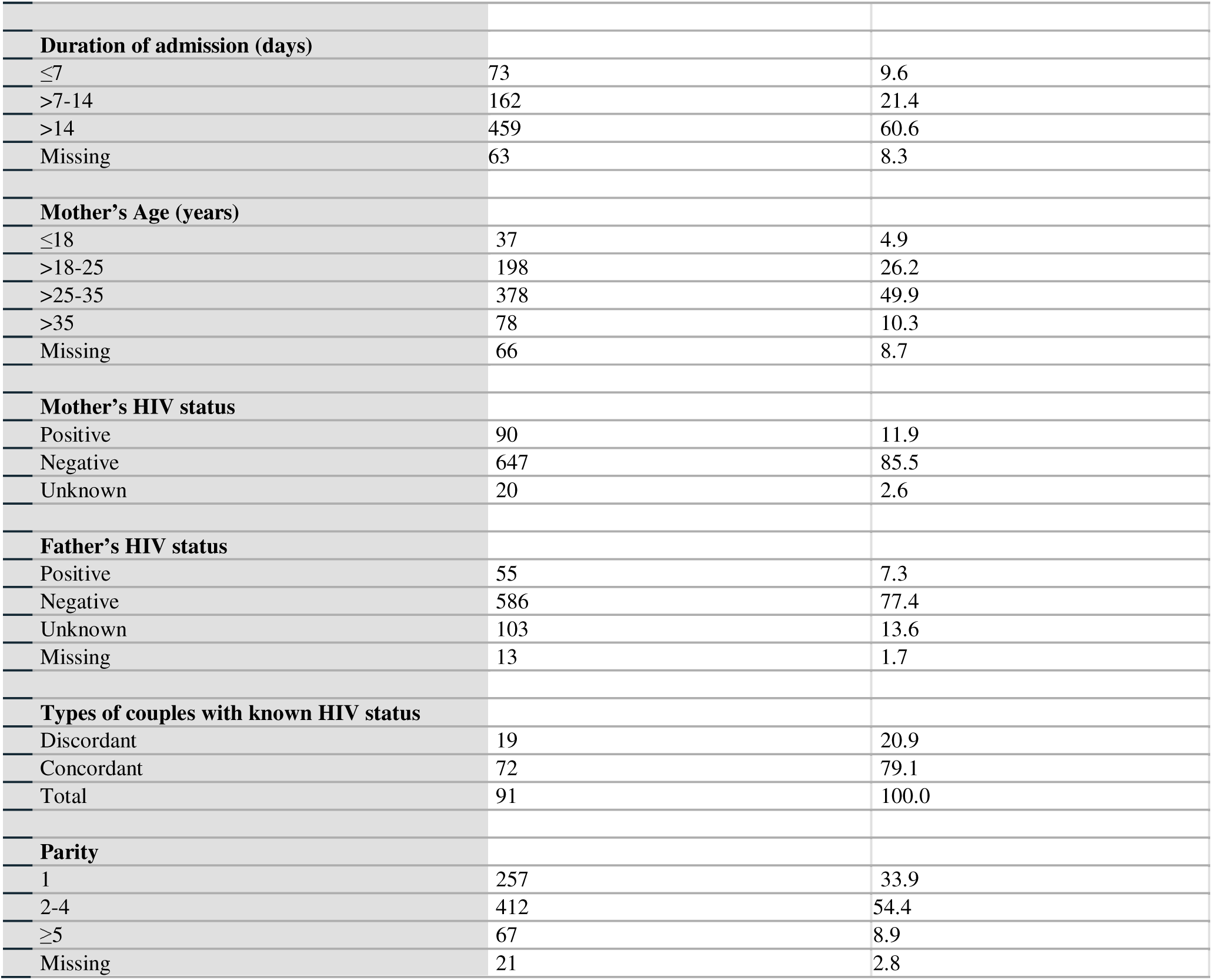
Characteristics of Mother-neonate participants in the study, n=757.

Mothers and Fathers tested positive for HIV at rates of 11.9% (90 out of 757) and 7.3% (55 out of 757), respectively. It is worth noting that Fathers had a higher number and percentage of unknown HIV status (103, 13.6%) than mothers (20, 2.6%). As a result, 51.7% of mothers transmitted HIV to their children, while 48.3% of infants were exposed to HIV. Four mothers (0.5%) had previously lost one to three pregnancies. Additionally, 29 (3.8%) mothers had no documented prenatal care during their pregnancy.

The incidence of adolescent pregnancy was 4.9% (37 out of 757); the overall Cesarean section rate was 20.6% (156 out of 757); 434 (57.3%) delayed the initiation of breastfeeding; the majority were referred from other health facilities (622, 82.2%); 543 (73.6%) neonates showed growth faltering; and a higher percentage presented at admission with abnormal body temperatures (60.1%), heart rates (66.8%), and respiratory rates (83%). Table 1 shows that there were some missing records for the variables in the ward registration and files.

### Characteristics of HIV positive vs negative neonates in the study

Bivariate analysis was performed to investigate the relationships between the independent variables and neonatal HIV status, Table 2. Data belonging to 52 Neonates with unknown HIV status were further removed from this section of analysis.

**Table 2:** Crosstabulation of independent variables with known neonatal HIV status in the study, n=705.

|  |  | Infant HIV status |  | Total |
| --- | --- | --- | --- | --- |
|  |  | Positive (%) | Negative (%) |  |
| **Sex** | Chi=0.188 p-value=0.665 |  |  |  |
| Male | OR 0.85 (95% CI 0.42, 1.75) | 16 (51.6) | 374 (55.6) | 390 (55.4) |
| Female |  | 15 (48.4) | 299 (44.4) | 314 (44.6) |
| **Total** |  | 31 | 673 | 704 |
| **Body of neonate temperature at admission** | Fisher's Exact Test =1.000 |  |  |  |
| Hypo (<36.5°C)/Hyperthermia (>37.5 °C) | OR=1.05. 95% CI 0.47, 2.34 | 14 (56.0) | 335 (54.9) | 349 (55.0) |
| Normal (36.5.-37.5°C) |  | 11 (44.0) | 275 (45.1) | 286 (45.0) |
| **Total** |  | 25 | 610 | 635 |
| **Heart rate at admission** | Chi=0.793 p-value=0.373 |  |  |  |
| Brady/Tachycardia | OR 1.51 (95% CI 0.61, 3.76) | 15 (68.2) | 325 (58.7) | 340 (59.0) |
| Normal Heart rate |  | 7 (31.8) | 229 (41.3) | 236 (41.0) |
| **Total** |  | 22 | 554 | 576 |
| **Resp rate at admission** | Chi=0.319 p-value=0.572 |  |  |  |
| Brady/Tachypnea | OR 1.37 (95% CI 0.46, 4.11) | 19 (82.6) | 402 (77.6) | 421 (77.8) |
| Normal Resp rate |  | 4 (17.4) | 116 (22.4) | 120 (22.2) |
| **Total** |  | 23 | 518 | 541 |
| **Postnatal growth** | Chi=6.674 p-value=0.010 |  |  |  |
| Faltering | OR 3.02 (95% CI 1.26, 7.24) | 20 (74.1) | 290 (48.7) | 310 (49.8) |
| Thriving |  | 7 (25.9) | 306 (51.3) | 313 (50.2) |
| **Total** |  | 27 | 596 | 623 |
| **Syphilis status of Neonate** | Fisher's Exact Test= 0.093 |  |  |  |
| Positive | OR 2.61 (95% CI 0.87, 7.86) | 4 (12.9) | 36 (5.4) | 40 (5.7) |
| Negative |  | 27 (87.1) | 634 (94.6) | 661 (94.3) |
| **Total** |  | 31 | 670 | 701 |
| **Gestational age** | Fisher's Exact Test= 0.047 |  |  |  |
| Preterm | OR =5.83 (95%CI 1.15, 29.58) | 3 (50.0) | 58 (14.6) | 61 (15.2) |
| Term |  | 3 (50.0) | 338 (85.4) | 341 (84.8) |
| **Total** |  | 6 | 396 | 402 |
| **Mode of Delivery** | Chi=0.044<br>p.value=0.834 |  |  |  |
| Cesarean section | OR=0.907. 95% CI 0.37, 2.25 | 6 (19.4) | 141 (20.9) | 147 (20.9) |
| Vaginal Delivery |  | 25 (80.6) | 533 (79.1) | 558 (79.1) |
| **Total** |  | 31 | 674 | 705 |
| **Birthweight status** | Chi= 0.141 p.value= 0.707 |  |  |  |
| Low birthweight | OR=1.19. 95% CI 0.48, 2.98 | 6 (20.0) | 112 (17.3) | 118 (17.5) |
| Normal/High birthweight |  | 24 (80.0) | 534 (82.7) | 558 (82.5) |
| **Total** |  | 30 | 646 | 676 |
| **Time of initiating BF** | Chi=1.045 p.value=0.307 |  |  |  |
| Delayed (>1hour) | OR=0.69. 95% CI 0.33, 1.42 | 15 (48.4) | 383 (57.7) | 398 (57.3) |
| Early (≤1hour) |  | 16 (51.6) | 281 (42.3) | 297 (42.7) |
| **Total** |  | 31 | 664 | 695 |
| **Any ANC** | Fisher's Exact Test =1.000 |  |  |  |
| No | OR=1.04. 95% CI 0.14, 7.97 | 1 (3.2) | 21 (3.1) | 22 (3.1) |
| Yes |  | 30 (96.8) | 653 (96.9) | 683 (96.9) |
| **Total** |  | 31 | 674 | 705 |
| **Referral** | Chi=0.078 p=0.780 |  |  |  |
| Hospital Facility | OR=1.15. 95% CI 0.43, 3.05 | 26(83.9) | 552 (81.9) | 578 (82.0) |
| Self |  | 5 (16.1) | 122 (18.1) | 127 (18.0) |
| **Total** |  | 31 | 674 | 705 |
| **Parity (# of deliveries)** | Chi 4.577 p=0.032 |  |  |  |
| Primip(1)/ Multiparous (≥5) | OR=0.399. 95% CI 0.17, 0.95 | 7 (25.0) | 300 (45.5) | 307(44.7) |
| Moderate Parity (2-4) |  | 21 (75.0) | 359 (54.5) | 380 (55.3) |
| **Total** |  | 28 | 659 | 687 |
| **Place of Delivery** | Chi= 0.000 p=0.985 |  |  |  |
| Outside Health Facility | OR=0.986. 95% CI 0.23, 4.27 | 2 (6.5) | 44 (6.5) | 46 (6.5) |
| Health Facility |  | 29 (93.5) | 629 (93.5) | 658 (93.5) |
| **Total** |  | 31 | 673 | 704 |
| **Type of couples** | Fisher's Exact Test =0.001 |  |  |  |
| Discordant | OR=11.72. 95% CI 3.35, 40.99 | 4 (15.4) | 9 (1.5) | 13 (2.1) |
| Concordant |  | 22 (84.6) | 580 (98.5) | 602 (97.9) |
| **Total** |  | 26 | 589 | 615 |

The profile of the HIV positive neonates were similar in almost all aspects investigated, to those of the HIV negative neonates, except that: the HIV positive neonates were more likely to be from discordant couples (OR=11.72. 95% CI 3.35, 40.99; Fisher=0.001); mothers of moderate parity (OR=2.51. 95% CI 1.05, 5.88; p=0.032); they were more likely to be born premature OR 5.83 (95%CI 1.15, 29.58; p=0.047) and their postnatal growth was significantly deficient OR 3.02 (95% CI 1.26, 7.24; p=0.010) compared to those who were HIV negative, Table 2.

#### Frequency of antenatal care

One-way ANOVA Test was performed to compare the means of the frequency of ANC visits between mothers of neonates with HIV positive, negative and unknown statuses in the study. Overall mean frequency of ANC visits was 5.28 (2.09). The mean frequency of ANC visits of the mothers with 31 HIV positive neonates was 5.39 (sd 2.09), that of the 674 HIV negative neonates was 5.32 (sd 2.03) and that of the 52 unknown HIV status neonates was 4.60 (sd 2.7). Levene statistics was significant at p-value of 0.021, hence unequal variances was assumed. The mean difference of the frequency of ANC visits between the 3 groups was statistically insignificant, p=0.051, even though those with unknown HIV status was the lowest of the 3 groups.

#### Neonate’s chronological age at admission

The data for chronological age at admission was not normally distributed (skewness 3.10, Kurtosis 22.13). Hence, Kruskal Wallis’ test was performed to compare the median age at admission between HIV positive, negative and unknown status neonates in the study. Overall median age was 14days (IQR 9, 21). The median age at admission of the 24 HIV positive neonates was 15days ([IQR 8.25, 24], that of the 646 HIV negative neonates was 14 days (IQR 9, 21) and those of the 51 neonates with unknown HIV status was 17days (IQR 9, 23). The difference in the median age at admission between the 3 groups was statistically insignificant, p=0.508, even though the neonates with unknown HIV status tend to present later than the rest of the group.

#### Random Blood Sugar (RBS) at admission

One-way ANOVA Test was performed to compare the means of RBS at admission between neonates with HIV positive, negative and unknown statuses in the study. Overall mean RBS was 5.36mmol/l (sd 1.18). The mean RBS of 27 HIV positive neonates was 5.21mmol/l (sd 1.129), that of the 585 HIV negative neonates was 5.33mmol/l (sd 1.101) and that of the 41 unknown HIV status neonates was 5.82 (sd 1.97). Levene statistics was significant at p-value of 0.007, hence unequal variances was assumed. The difference in the mean RBS between the 3 groups was statistically significant, p=0.029, and those with unknown HIV status had the highest RBS of the 3 groups.

#### Oxygen saturation (O2) at admission

One-way ANOVA Test was performed to compare the means of O2 at admission between neonates with HIV positive, negative and unknown statuses in the study. Overall mean O2 was 96.6% (sd 3.174). The mean O2 of the 26 HIV positive neonates was 96.58% (sd 2.176), that of the 611 HIV negative neonates was 96.68% (sd 3.252) and that for the 45 unknown HIV status neonates was 96.4% (sd 2.55). Levene test was insignificant at p-value of 0.502, hence equal variances was assumed. The mean O2 differences between the 3 groups were statistically insignificant, p=0.845.

#### Maternal age

One-way ANOVA Test was performed to compare the mean maternal age between neonates with HIV positive, negative and unknown statuses in the study. The mean maternal age of the 28 HIV positive neonates was 31.1years (sd 5.40), that of the 626 HIV negative neonates was 27.7years (sd 5.76) and that for the 37 unknown HIV status neonates was 30.49years (sd 5.82). Levene test was insignificant at p-value of 0.928, hence equal variances was assumed. The difference in mean maternal age across the 3 groups was statistically significant, p<0.001, with mothers of HIV negative mothers being the youngest.

#### Duration of admission

The data for duration of admission was not normally distributed (skewness 2.19, Kurtosis 7.6). Hence, Kruskal Wallis’ test was performed to compare the median duration of admission between HIV positive, negative and unknown status neonates in the study. Overall median duration of admission was 6days (IQR 4, 9). The median duration of admission of the 24 HIV positive neonates was 6days ([IQR 4,8], that of the 646 HIV negative neonates was 6 days (IQR 4, 9) and those of the 46 neonates with unknown HIV status was 7days (IQR 5, 10). The difference in the median duration of admission between the 3 groups was statistically insignificant, p=0.559, even though the neonates with unknown HIV status tend to stay longer on the ward than the rest of the group.

### Characteristics of neonates with unknown vs known HIV status in the study

Fifty-two neonates had unknown HIV status. Majority of these neonates with unknown HIV status were female (27, 51.9); term gestation (26, 50%); delivered at health facilities (51, 98.1%) and vaginally (43, 82.7%); delayed in initiating breastfeeding (36, 69.2%); were more than 7days old (42, 80.8%) at admission; had mothers aged 25-35years, (18, 34.6%); of moderate parity (32, 61.5%); had mothers who were HIV positive (30, 57.7%), but with Fathers whose HIV status were unknown (27, 51.9%); referred from other health facilities (44, 84.6%); abnormal heart rates (19, 36.5%), respiratory rates (28,53.8%) and temperatures (28, 53.8%), normal birth weight (38, 73.1%), oxygen saturation (38, 73.1%) and random blood sugar (38, 73.1%) at admission; negative syphilis status (47, 90.4%) and were hospitalized for less than 7days (27, 51.9%). Discordant couples rate in general was 20.9% (19/91). Of note, 6/22 (27.3%) of infants with unknown HIV belonged to discordant parents, which was the highest rate compared to those with positive HIV status (4/26, 15.4%) and negative HIV status (9/589, 1.5%), p<0.001. In all of the 19 discordant couples, 18 of the Mothers were positive, while only 1 Father was positive.

When compared with infants, whose HIV status was known, infants with unknown HIV status were significantly more likely to have received no antenatal care (p<0.001; OR=4.83. 95% CI 1.96, 11.91); belonged to parents with unknown or positive HIV status, p<0.001 or discordant parents (p<0.001; 10/50, 20%; OR=9.07; 95% CI 3.16, 26.00) and elderly mothers (p=0.035; OR=2.94. 95% CI 1.15, 7.51). These infants with unknown HIV status were also 1.94 times likely to be born premature (p=0.103), 1.68 times (p=0.092) more likely to have delayed in initiating breastfeeding from birth, 1.66 times (p=0.118) more likely to be growth faltering and 1.50 times (p=0.218) more likely to present with abnormal body temperatures compared to infants with known HIV status in the study, Table 3.

**Table 3:**
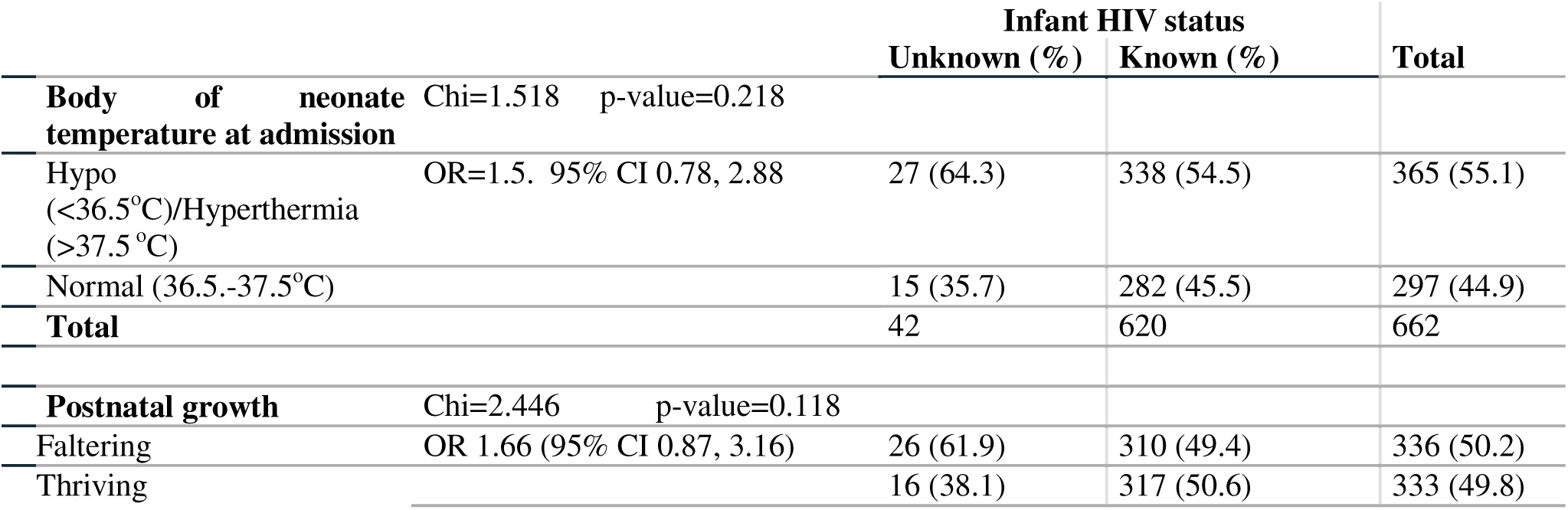
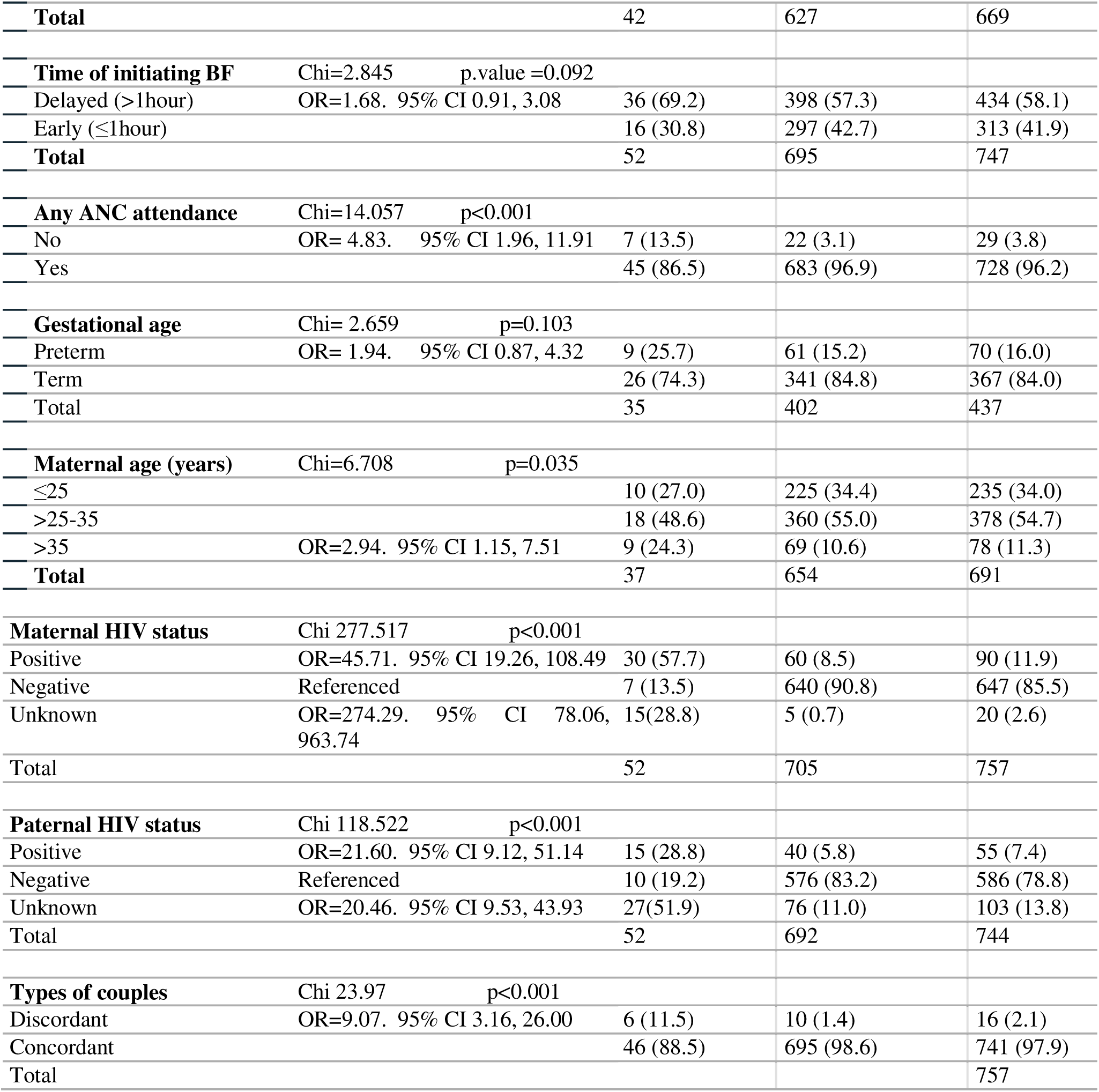
Characteristics of neonates with unknown vs known HIV positive status in the study, n=757.

|  |  | Infant HIV status |  | Total |
| --- | --- | --- | --- | --- |
|  |  | Unknown (%) | Known (%) |  |
| **Body of neonate temperature at admission** | Chi=1.518 p-value=0.218 |  |  |  |
| Hypo (<36.5°C)/Hyperthermia (>37.5 °C) | OR=1.5. 95% CI 0.78, 2.88 | 27 (64.3) | 338 (54.5) | 365 (55.1) |
| Normal (36.5.-37.5°C) |  | 15 (35.7) | 282 (45.5) | 297 (44.9) |
| **Total** |  | 42 | 620 | 662 |
| **Postnatal growth** | Chi=2.446 p-value=0.118 |  |  |  |
| Faltering | OR 1.66 (95% CI 0.87, 3.16) | 26 (61.9) | 310 (49.4) | 336 (50.2) |
| Thriving |  | 16 (38.1) | 317 (50.6) | 333 (49.8) |
| **Total** |  | 42 | 627 | 669 |
| **Time of initiating BF** | Chi=2.845 p.value =0.092 |  |  |  |
| Delayed (>1hour) | OR=1.68. 95% CI 0.91, 3.08 | 36 (69.2) | 398 (57.3) | 434 (58.1) |
| Early (≤1hour) |  | 16 (30.8) | 297 (42.7) | 313 (41.9) |
| **Total** |  | 52 | 695 | 747 |
| **Any ANC attendance** | Chi=14.057 p<0.001 |  |  |  |
| No | OR= 4.83. 95% CI 1.96, 11.91 | 7 (13.5) | 22 (3.1) | 29 (3.8) |
| Yes |  | 45 (86.5) | 683 (96.9) | 728 (96.2) |
| **Gestational age** | Chi= 2.659 p=0.103 |  |  |  |
| Preterm | OR= 1.94. 95% CI 0.87, 4.32 | 9 (25.7) | 61 (15.2) | 70 (16.0) |
| Term |  | 26 (74.3) | 341 (84.8) | 367 (84.0) |
| **Total** |  | 35 | 402 | 437 |
| **Maternal age (years)** | Chi=6.708 p=0.035 |  |  |  |
| ≤25 |  | 10 (27.0) | 225 (34.4) | 235 (34.0) |
| >25-35 |  | 18 (48.6) | 360 (55.0) | 378 (54.7) |
| >35 | OR=2.94. 95% CI 1.15, 7.51 | 9 (24.3) | 69 (10.6) | 78 (11.3) |
| **Total** |  | 37 | 654 | 691 |
| **Maternal HIV status** | Chi 277.517 p<0.001 |  |  |  |
| Positive | OR=45.71. 95% CI 19.26, 108.49 | 30 (57.7) | 60 (8.5) | 90 (11.9) |
| Negative | Referenced | 7 (13.5) | 640 (90.8) | 647 (85.5) |
| Unknown | OR=274.29. 95% CI 78.06, 963.74 | 15(28.8) | 5 (0.7) | 20 (2.6) |
| **Total** |  | 52 | 705 | 757 |
| **Paternal HIV status** | Chi 118.522 p<0.001 |  |  |  |
| Positive | OR=21.60. 95% CI 9.12, 51.14 | 15 (28.8) | 40 (5.8) | 55 (7.4) |
| Negative | Referenced | 10 (19.2) | 576 (83.2) | 586 (78.8) |
| Unknown | OR=20.46. 95% CI 9.53, 43.93 | 27(51.9) | 76 (11.0) | 103 (13.8) |
| **Total** |  | 52 | 692 | 744 |
| **Types of couples** | Chi 23.97 p<0.001 |  |  |  |
| Discordant | OR=9.07. 95% CI 3.16, 26.00 | 6 (11.5) | 10 (1.4) | 16 (2.1) |
| Concordant |  | 46 (88.5) | 695 (98.6) | 741 (97.9) |
| **Total** |  |  |  | 757 |

## Discussion

This investigation recorded the human immunodeficiency virus (HIV) status of infants admitted to the NCOE at UTH-Children Hospital throughout a twelve-month duration. Overall, the 2024 annual prevalence of HIV and syphilis among all neonates admitted was observed to be 4.1% and 5.9%^8^, respectively. The HIV status remained unknown for 52 admitted neonates, representing 6.9% of the total. The prevalence of HIV among mothers was 11.9%, while among fathers, it was 7.3%. Notably, a greater proportion and number of fathers had an unknown HIV status (13.6%, 103 individuals) compared to mothers (2.6%, 20 individuals). Consequently, the rate of mother-to-child HIV transmission was observed to be 51.7%, with an infant HIV exposure rate of 48.3%. The incidence of discordant couples was 20.9%. Furthermore, 3.8% of mothers did not attend antenatal clinic visits during gestation. Teenage pregnancy accounted for 4.9% of cases, and 6.2% of deliveries occurred outside of a healthcare facility. The cesarean section rate was 20.9%. Delayed initiation of breastfeeding was reported in 57.3% of cases. A significant majority, 82.2%, were referrals from other healthcare facilities. Growth faltering was present in 73.6% of infants, and a higher percentage of neonates presented upon admission with abnormal body temperatures (60.1%), heart rates (66.8%), and respiratory rates (83%). A frequent challenge encountered was the absence of records within the ward register and associated files.

The demographic and clinical profiles of neonates testing positive for HIV were found to be comparable across all evaluated parameters to those of HIV-negative neonates. However, a notable distinction was the significantly older average maternal age (31.1 years) among mothers of HIV-positive infants. These mothers also exhibited a greater propensity for moderate parity, were more likely to have given birth to infants from discordant couples (OR 11.72), and their infants were more prone to prematurity. Furthermore, the postnatal growth of HIV-positive infants was demonstrably impaired compared to their HIV-negative counterparts.

The 52 infants whose HIV status remained unknown presented with distinct characteristics compared to those with identified HIV status. Specifically, these neonates with unknown HIV status were more frequently born to parents with positive or unknown HIV status, or to sero-discordant couples, as opposed to HIV-negative parents. They also did not receive antenatal care, experienced delays in commencing breastfeeding, exhibited growth faltering, and presented with relatively elevated random blood glucose levels and abnormal body temperatures upon admission. All mothers and infants with a confirmed positive HIV status were undergoing antiretroviral (ARV) therapy, and exposed infants were administered ARV prophylaxis. Nevertheless, the precise commencement dates of these treatments or prophylactic measures could not be ascertained from the available records.

### Prevalence of HIV

The infant HIV incidence of 4.1% observed in this investigation is lower than the 5.5% reported by Kassie et al. in 2020^9^, who conducted a retrospective chart review of 239 infants at a specialized hospital in Northwestern Ethiopia. A descriptive observational study by Torpey et al. in 2012^10^ examining HIV Early Infant Diagnostic activities across five Zambian provinces from September 2007 to July 2010 recorded an HIV prevalence of 7.1% in infants aged 0 to 6 weeks. Higher prevalence rates were documented in Northwest Ethiopia at 8.1% (Tiruneh et al., 2022)^11^ and in Ghana at 11.1% (Ahenkan et al., 2025)^12^, both exceeding the 4.1% identified in the present study. In contrast, countries such as Tanzania (0.4%) and Eswatini (2.7%) have significantly lower prevalence rates than those found in this study, as noted by Lutz in 2022^13^. Since 2015, the Pan American Health Organisation (PAHO)/World Health Organization (WHO)^14^ has officially recognized Cuba’s achievement in eradicating mother-to-child HIV transmission. The maternal HIV rate of 11.9% recorded in this study is double the 5.1% reported in Rwanda by Ogah et al. in 2023^15^. Data from ZAMPHIA (2022)^16^ indicated maternal HIV rates of 13.9% and paternal HIV rates of 8%, both of which were higher than the maternal rate of 11.9% and paternal rate of 7.3% observed in the current research. In terms of trends, the comparatively low rates of maternal, paternal, and neonatal HIV positive rates recorded in this study reflect the well-organized and effective PMTCT program in the nation of Zambia.

### Uptake of early infant HIV testing/Discordant couples

Hampanda et al. (2017)^17,18^ observed that only 72.8% of infants were tested for HIV at 4-6 weeks of age among 320 HIV-positive mothers who brought their children in for regular pediatric vaccinations at a major public health facility in Lusaka, Zambia, during a cross-sectional study. Likewise, according to data from UNAIDS (2020)^19^, at least 30% of infants exposed to HIV in Zambia do not receive Early Infant Diagnosis (EID) testing within the first two months after birth; this figure is 24.5% in Tanzania (Kiurugo et al., 2024)^20^. The proportion of infants with unknown HIV status in earlier studies was approximately four times higher than the 6.9% noted in this current research, further demonstrating the advancements in PMTCT services within Zambia. Fathers were more prone to have unknown HIV status. Various obstacles within healthcare systems hinder the early detection of HIV infection in infants in low-resource environments. For instance, DNA-PCR using dried blood spots demands advanced and costly nucleic acid testing, along with highly skilled personnel and sophisticated lab infrastructure, which is typically found only in large, centralized labs. In fact, in Zambia, there exist only three facilities with the necessary laboratory capabilities to conduct PCR (ZMOH, 2010)^18^. This problem is further compounded by limitations on sample collection locations to PMTCT clinics, shortages of HIV testing supplies, ineffective sample transportation systems, and challenges in supply chain management. Limiting HIV testing to specific locations and office hours may cause some babies who arrive outside these times, including weekends, to be missed.

Furthermore, social factors such as emotional intimate partner violence directed at females, the controlling behavior of male partners (Simons-Morton, 2009)^21^, awareness of HIV status prior to pregnancy, disclosure of HIV status to partners, adherence to antiretroviral therapy (ART), and having a partner with a known HIV status significantly affect the rates of early infant testing. A major issue highlighted in this study is that most infants with unknown HIV status were from parents whose HIV status was either positive, unknown, or sero-discordant. Social factors such as disclosure issues may have contributed to the high rate of unknown newborn HIV status in this study, as the discordant couples rate was 20.9% overall, but highest at 27.3% among infants with unknown HIV status as against 20% among infants with known HIV status. Zambia has a discordant couples rate of 20% (21% in Lusaka), which is congruent with estimates from Uganda, Rwanda, Tanzania, and Kenya (Lambdin et al., 2021)^22^ and similar to what was observed in this study.

Another possible cause for unknown HIV status is the question of consent, which is required as part of the country’s formal HIV testing method. Despite proper counselling, some parents refuse to test themselves or their newborn for fear of the psychological impact and stigma associated with receiving negative news. Delays in determining the infant’s HIV status will postpone the commencement of treatment or prophylaxis, perpetuating the cycle of rising HIV incidence in the community, and leading to undesirable consequences.

Notably, the HIV-positive neonates were 11.72 times (15.4%) more likely to be born to discordant couples than HIV-negative neonates (1.5%). Infants with unknown HIV status have demographic and clinical characteristics similar to HIV-positive infants, such as sero-discordant parental partnerships, elderly motherhood and growth abnormalities. Consequently, it would be a regrettable outcome if these infants were denied the opportunity to undergo PCR testing and possibly begin antiretroviral therapy, if confirmed positive during their hospital stay.

To address this problem of low uptake of HIV testing, especially among male partners, the PMTCT program made it crucial for fathers to participate and support these efforts. The program also ensures that information is shared with men and that discussions occur in venues beyond the clinics frequented by pregnant women, since those clinics are typically designed for women and not always conducive to men’s involvement (Kankasa et al., 2002). These strategies include targeting male community leaders with PMTCT education and information, providing community education on PMTCT in venues where men gather, such as bars, football fields, and taxi stands and establishing discussion and support groups for men, (Kankasa et al., 2002; Sinkala,et al. 2011)^3,23^.

### HIV MCT rate

Zambia’s mother-to-child HIV transmission (MCT) rate has drastically fallen to an anticipated 5.9% (at the national level) by 2025, down from about 30% in 2005, 24% in 2009, and 12% in 2012. These rates remain above the worldwide goal rate of <5%. The decline in MCT rates in Zambia over the last two decades can be attributed to widespread ART treatment among more than 90% of HIV-positive pregnant mothers, with coverage reaching 100% in some areas such as Kitwe (National HIV/AIDS/STI/TB Council, 2025)^24^. This highlights the need of knowing one’s HIV status. However, in this current study, at the NCOE, UTH-Children, the HIV MCT rate was extremely high, at 51.7%. This demonstrates that, despite MTCT prevention initiatives, transmission rates remain high, similar to the pre-intervention era. However, keep in mind that the study only looked at unwell neonates who needed to be admitted to a national referral hospital, and data on mothers and infants with unknown HIV status was excluded from the analysis.

### Profile of HIV positive infants

The similarity in characteristics of HIV positive infants in this study to HIV negative infants, especially when comparing their intrauterine growths, could be ascribed to the efficiency of Zambia’s PMTCT programs. The areas of dissimilarity such as elderly motherhood, moderate parity, preterm birth, and postnatal growth deficiency (strongly associated with HIV positive infants) may indicate targeted areas for future interventions and improved infant morbidity and mortality.

The age distribution of HIV positive mothers in this study (60.7% were between the ages of 25 and 35, and 60.7% of HIV positive infants were in this age group) is consistent with the findings in Kassie et al. (2020)^9^, who found that 77.1% of mothers of HIV positive infants were between the ages of 25 and 35. Unlike the preterm delivery reported by Benali et al. (2019)^25^ and our current investigation, Kassie et al (2020)^9^. found that post-term birth was related with HIV positive.

### Antenatal records/Unknown HIV status

In Benali et al’s research (2019)^25^, which included 5029 newborn admissions at Charlotte Maxeke Johannesburg Academic Hospital in South Africa in a cross-sectional study between 2015 and 2017, 75% of the mothers attended ANC, which is lower than the 96.2% observed in this present study. Unexpectedly, there was no significant difference in ANC attendance and frequency of visits between mothers of HIV PCR positive (96.8%) and HIV PCR negative (96.9%) newborns. However, mothers of neonates with unknown HIV status were 2.94 times more likely to be elderly, nearly 5 times more unlikely to visit prenatal clinics, and even if they do, their attendance was irregular when compared to those with known HIV status. Despite efforts by the government and other organizations in the country to educate women, this data demonstrates that a significant percentage of mothers are still unaware of the importance of attending ANCs and testing for HIV early.

## Postnatal growth

It is worth noting that there was no significant difference in intrauterine growth between all of the neonates in the study (HIV positive, negative, and unknown status), as indicated by the assessment of their birthweights-, indicating an effective PMTCT program. However, changes in growth pattern became obvious shortly after birth, much earlier than the 3-4 month age reported by Isanaka et al. (2009)^26,27,28^. Furthermore, the growth faltering rate among HIV positive neonates in this study (74.1%) was higher than the 20-70% quoted in Isanaka et al’s systematic evaluation of 37 papers on HIV infection and postnatal growth. The rate of growth faltering among HIV-positive infants in this study was also significantly higher than that reported by Kartik et al. (2010)^29^ in a cohort study of 840 mother-infant pairs in South Africa. Several mechanisms have been proposed to explain how HIV impacts growth. These include the direct effect of the HIV virus on normal cellular processes, leading to dysregulation; weakening of the immune system, which makes infants more vulnerable to recurrent infections such as diarrhea, pneumonia, and tuberculosis that interfere with nutrient absorption and utilization; alterations in metabolism caused by HIV and its treatments, affecting fat distribution and nutrient processing; metabolic side effects from some antiretroviral drugs, despite their essential role; and a strong association between higher viral loads and poorer growth, particularly during the first months of life. This report suggests gaps in postnatal growth monitoring and care of these infants that requires intervention.

## Limitations

Incomplete records being a retrospective study.

## Strength

Large sample sized data complied over a long period of time.

## Conclusion

The significant number of mothers lacking antenatal care in this study is a cause for concern and poses a risk to the advancement of the Prevention of Mother-to-Child Transmission (PMTCT) program within the nation. Intensified antenatal care initiatives, encompassing early HIV screening for expectant mothers and their partners, are imperative to enable the timely initiation of Antiretroviral Therapy (ART) and PMTCT services. Particular vigilance is warranted for neonates presenting with an unknown HIV status in healthcare facilities within resource-limited environments. Such infants, especially those born to elderly mothers with inadequate or absent antenatal care, those with moderate parity, premature births, delayed breastfeeding initiation, faltering growth, and abnormal vital signs, should be suspected of HIV positivity and undergo early infant testing to further mitigate infant morbidity and mortality. Regular assessment of these infants’ feeding, health, and growth shortly after birth is crucial, either through home visits or postnatal clinic appointments. Targeted counseling for mothers (and partners) with unknown HIV status, HIV positive infants, infants with unknown HIV status, women aged >25 years, and discordant couples; and public education are essential for reducing HIV incidence and improving infant health outcomes within the community.

## Data Availability

All data produced in the present study are available upon reasonable request to the authors

## Author Contributions

The corresponding author (Dr Adenike Oluwakemi Ogah) generated the study title and concept, collected the raw data, conducted the data analysis, interpreted the results and drafted this manuscript. Dr James-Aaron Ogbole Ogah co-managed the data and assisted in the drafting of the manuscript. Dr Edwin Kanombola Chembo entered and managed the data. All the authors contributed to the intellectual content of this manuscript and final editing of the article.

## Acknowledgements

The authors are extremely grateful to the participants involved in this study, to the NCOE staff of UTH-Children Hospital in Lusaka that contributed to this study and to the research team.

